# Algorithm-Based Model for Gastrointestinal and Liver Histopathological Analysis Using VGG16 and Specialized Stains: Statistical Validation of Thresholds in AI-Driven Digital Pathology

**DOI:** 10.64898/2026.04.08.26350456

**Authors:** Adekunle O. Adeluwoye, Majesty O. Gbadegesin, Favour M. James, Peter S. Otegbade, Ayanfeoluwa Alabetutu

**Author notes:** Corresponding author: Adekunle O. Adeluwoye.

## Abstract

Digital pathology, coupled with advanced image recognition algorithms, represents a transformative frontier in histopathological diagnosis. This sub-Saharan African laboratory’s exploratory study investigates the application of a Convolutional Neural Network (CNN) model, specifically leveraging the VGG16 architecture with transfer learning, for automated analysis and classification of selected gastrointestinal (GIT) and liver tissue samples, incorporating both routine and specialized staining protocols. The study utilized a dataset comprising 114 samples (18 liver, 96 GIT images) derived from archival formalin-fixed paraffin-embedded tissue blocks at University College Hospital, Ibadan, Nigeria. Specialized staining techniques included Alcian Yellow for GIT mucin visualization and Masson’s Trichrome for liver fibrosis assessment, alongside conventional H&E staining. Model performance was evaluated using statistical methodologies including Wilson Score confidence intervals (CI), Bayesian probability assessment, and effect size analysis. Results reveal a striking dichotomy in model performance. The GIT tissue model achieved perfect classification accuracy (100% test accuracy) with exceptional statistical significance (Z=10.0, p<0.0001), Wilson CI [96.29%, 99.99%], Cohen’s h=1.571, and Bayesian probability >99.99%. Conversely, the liver tissue model demonstrated diagnostic failure (42.86% test accuracy), with Z=-1.428, p=0.9236, Wilson CI [33.59%, 52.65%], Cohen’s h=-0.144, and Bayesian probability of 7.64%. This performance divergence correlates with training data availability, as the liver dataset fell far below empirically established thresholds (>100–200 samples) for reliable classification. The liver model’s failure reveals limitations in transfer learning with insufficient data. These findings underscore critical implications for AI-enhanced digital pathology, demonstrating potential deployment of the GIT model as a promising one that supports tissue-specific model development.

## 1. Introduction

The accurate and timely diagnosis of diseases affecting the gastrointestinal tract and the liver remains a cornerstone of effective clinical management globally.^[1]^ These organ systems are frequently implicated in a spectrum of pathologies, ranging from chronic inflammatory conditions to aggressive malignancies, where early detection is paramount for favorable patient outcomes.^[2]^ Traditional histopathology, while maintaining its status as the gold standard for definitive diagnosis, is inherently reliant on the subjective interpretation of stained tissue sections by expert pathologists. This reliance introduces an element of inter-observer variability and can be a time-intensive process, particularly in high-volume settings or in regions facing a shortage of specialized personnel.^[3]^

The advent of digital pathology has provided a paradigm shift, moving the field from the analog microscope to a fully digitized workflow. This transformation involves the scanning of glass slides into high-resolution Whole Slide Images (WSIs), which then become amenable to computational analysis.^[1]^ This technological leap has paved the way for the integration of artificial intelligence (AI), particularly deep learning, to augment and potentially automate aspects of histopathological diagnosis. Among the various AI methodologies, Convolutional Neural Networks (CNNs) have emerged as the most potent tool for image recognition tasks in medicine, owing to their unique capacity to learn complex, hierarchical features directly from raw image data.^[4]^

In the context of histopathology, CNNs are adept at identifying subtle morphological changes that are indicative of disease, such as nuclear pleomorphism, architectural distortion, and the presence of specific cellular infiltrates. Previous studies have already highlighted the efficacy of AI in diagnosing various conditions, including hepatocellular carcinoma, chronic gastritis, and chronic hepatitis, with diagnostic accuracy often reported to be comparable to, or even exceeding, that of human pathologists.^[5, 6]^ Furthermore, the continuous refinement of segmentation techniques has significantly improved the automated identification of critical histological features, thereby enhancing the overall capability of automated pathology systems.^[7]^

This study focuses on a critical area of application: the automated classification of gastrointestinal and liver tissues using a VGG16-based CNN model. The rationale for selecting these tissues is multifold. Gastrointestinal diseases, such as inflammatory bowel disease and *Helicobacter pylori* infections, demand precise tissue-level characterization.^[2]^ Similarly, liver diseases, including cirrhosis and hepatocellular carcinoma, represent significant global health burdens that necessitate innovative diagnostic tools.^[8]^ Crucially, the study incorporates the analysis of tissues stained with specialized techniques—Alcian Yellow for mucin in GIT and Masson’s Trichrome for fibrosis in the liver—alongside the routine H&E. This approach is designed to validate the algorithm’s robustness in utilizing stain-specific features, which are often essential for definitive diagnosis in clinical practice. The use of transfer learning, by leveraging the pre-trained weights of the VGG16 model on the massive ImageNet dataset, is a pragmatic approach to address the often-limited size of medical image datasets, enabling the model to generalize effectively to the nuanced world of histopathology.^[9]^

Despite the evident promise, the deployment of AI in clinical pathology is not without its challenges. Issues such as the variability in staining protocols across different laboratories, inter-patient biological differences, and the ethical imperative for algorithmic transparency and explainability remain significant considerations.^[10, 11]^ By conducting this validation study within a high-volume clinical setting in a Sub-Saharan African context—the University College Hospital (UCH), Ibadan, Nigeria—the work also implicitly addresses the feasibility and generalizability of these advanced diagnostic tools in resource-constrained environments. The overarching goal is to demonstrate a robust, accurate, and clinically relevant AI-driven solution that can be seamlessly integrated into routine diagnostic workflows, thereby contributing to the global effort to improve diagnostic standards for liver and gastrointestinal pathologies.

## 2. Materials and Methods

### 2.1. Study Area and Design

This study employed an experimental, cross-sectional descriptive research design to validate the performance of image recognition algorithms in the automated analysis of histopathological images. The focus was on the application of CNNs for diagnosing pathologies in gastrointestinal and liver tissues, evaluating key performance metrics such as accuracy, sensitivity, specificity, and robustness. The work was conducted using archival formalin-fixed paraffin-embedded (FFPE) tissue blocks obtained from the Histopathology Laboratory at the University College Hospital (UCH), Ibadan, Nigeria, to help ensure a diverse and clinically relevant dataset. The samples included diseased and normal tissues, with GIT samples comprising cases of chronic gastritis, *H. pylori* infections, and inflammatory bowel diseases, and liver samples including hepatocellular carcinoma, chronic hepatitis, and cirrhosis. In this research, a dataset of 114 samples was utilized, comprising 18 images from liver biopsy tissue samples and 96 images from gastrointestinal tract (GIT) biopsy tissue samples. These images played a crucial role in training an machine learning (ML) model designed to recognize abnormal tissues within the liver and GIT. This design allowed for an in-depth assessment of the performance of machine learning models on a clinically relevant and diverse dataset. Only high-quality slides with confirmed expert diagnoses were included. Conversely, tissue sections exhibiting prominent staining artefacts or poor tissue integrity were excluded from the dataset.

### 2.2. Laboratory Procedures

#### 2.2.1. Sectioning

Tissue blocks were sectioned using a rotary microtome. The blocks were first trimmed and cooled for approximately 15 minutes to ensure consistent paraffin properties. They were then mounted, and sections were cut at a thickness of 5 micrometers. The resulting ribbon-like sections were floated on a water bath for flattening and subsequently collected onto slides.^[12]^

#### 2.2.2. Staining and Digital Imaging

In addition to the general tissue morphology provided by Hematoxylin and Eosin (H&E) staining, specialized stains were utilized to highlight specific features critical for diagnosis. Alcian Yellow was employed for the gastrointestinal tissues to identify mucin and gastric gland structures. Masson’s Trichrome was used for liver samples to highlight fibrosis and connective tissue, which is crucial for staging liver diseases such as cirrhosis. Digital imaging was performed using a high-resolution digital microscope with images saved in standardized formats (.tiff) for computational analysis.

### 2.3. Model Architecture and Training

The core of the image recognition task utilized a Convolutional Neural Network (CNN) based on the VGG16 architecture.^[13]^ VGG16 is a deep CNN, comprising 13 convolutional layers and three fully connected layers, known for its use of small 3×3 convolutional filters.^[14]^

The model was initialized using weights pre-trained on the ImageNet dataset through a process known as transfer learning, which significantly accelerates training and enhances generalization on smaller, specialized datasets.^[15]^ To optimize performance, several architectural modifications were incorporated. Max-pooling layers were strategically placed after groups of convolutional layers to reduce spatial dimensions and emphasize the most salient features. Batch normalization layers were introduced to stabilize the training process and enable the use of higher learning rates.^[16]^ To mitigate overfitting, dropout layers were added, randomly deactivating subsets of neurons during training and thereby encouraging the network to learn more robust and generalized features.^[17]^ The Rectified Linear Unit (ReLU) was employed as the activation function throughout the network due to its computational efficiency and ability to introduce non-linearity.^[18]^ Finally, the output layer utilized a softmax activation function to produce a probability distribution across the predefined classification categories, such as distinguishing between normal and abnormal tissue types for each organ.^[19]^

### 2.4. Dataset Preparation and Evaluation

The dataset was meticulously labeled according to tissue type and pathological condition. The machine learning model code processes an image, initially loading it from the specified path and performing various transformations. It rescales the image, adjusts its gamma for increased contrast, converts it to grayscale, and detects edges using the Canny edge detector. The results of each transformation are displayed side by side in a 1×4 grid using Matplotlib. Additionally, the output chart is saved as a PNG file in the specified directory. This comprehensive visualization allows for a detailed examination of the impact of each processing step on the original image, providing valuable insights into the image enhancement and feature extraction techniques applied.

Stratified sampling was employed to split the dataset into training and validation sets, ensuring that class distribution was maintained to prevent bias. Data augmentation techniques, including random rotations and shifts, were applied to the training set to enhance the model’s robustness and generalization capability. The final model performance was evaluated based on the testing accuracy on a held-out test set, as well as an analysis of training and validation loss and accuracy curves over epochs.

### 2.5. Reproducibility and Code Availability

To ensure reproducibility, all image preprocessing, model training, and evaluation steps were implemented in a Jupyter Notebook (.ipynb) environment. The notebook documents the full workflow, including gamma adjustment, grayscale conversion, edge detection, data augmentation, and CNN training parameters. Each code cell is annotated to allow replication of results and adaptation to new datasets. The notebook will be made available as supplementary material or via a public repository upon publication, enabling other researchers to reproduce our findings and extend the methodology to related histopathological tasks.

## 3. Results

### 3.1. Histological Feature Visualization

The initial study results involved the visualization of the processed tissue images. This included representations of the first four tissue micrographs for both gastrointestinal (Figure 3.1a) and liver tissue (Figure 3.1b), providing a baseline for the dataset’s visual characteristics.

**Figure: 3.1a:**
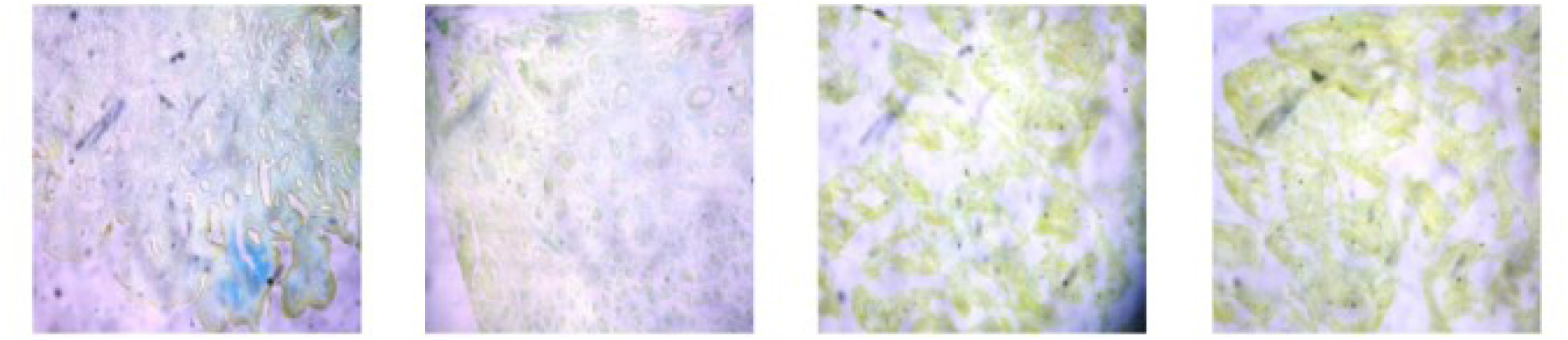
Visualization of the first 4 tissue pictures for the gastrointestinal tissue (Alcian Yellow stained slides)

Figure 3.1a and 3.1b showcases these images in a 1×4 grid, each presented without axis labels. This approach serves as a concise and effective means of visually inspecting and comparing staining patterns and tissue morphology across samples. The color variations aid in assessing structural features and staining consistency across the dataset, providing a convenient overview of the dataset.

**Figure: 3.1b:**
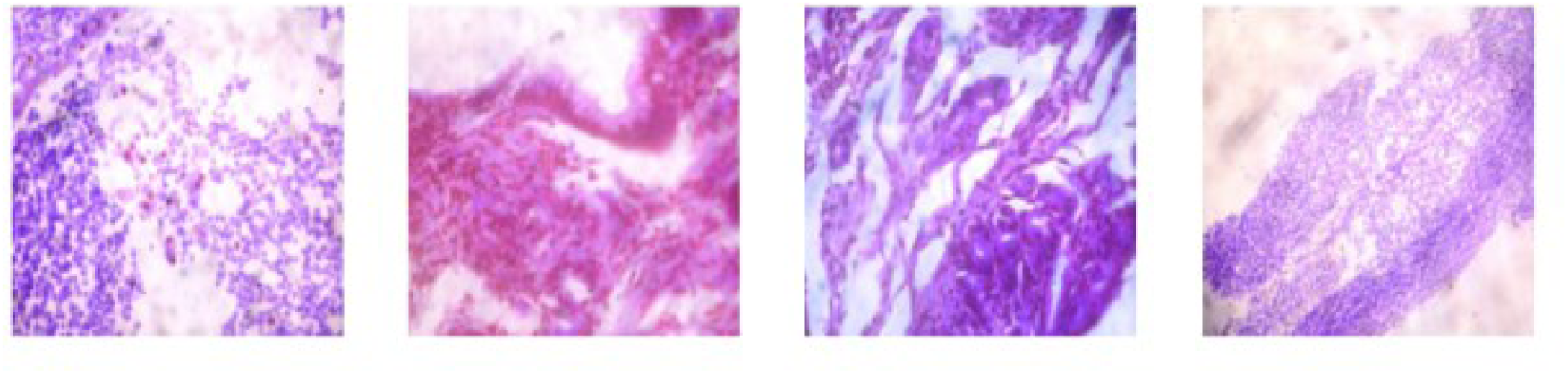
Visualization of the first 4 tissue pictures for the liver tissue (H&E stained slides)

### Visualization of abnormal and normal cell image

Furthermore, this work presented visualizations of both abnormal and normal tissue images.

Figures 3.2a - 3.2d demonstrates the impact of image processing techniques such as gamma adjustment, grayscale conversion, and canny edge detection on a single abnormal GIT tissue image stained with Alcian Yellow (Figure 3.2a), an abnormal GIT tissue image stained with H&E (Figure 3.2b) an abnormal liver tissue image stained with Masson Trichrome (Figure 3.2c), and an abnormal liver tissue image stained with H&E (Figure 3.2d).

**Figure 3.2a:**
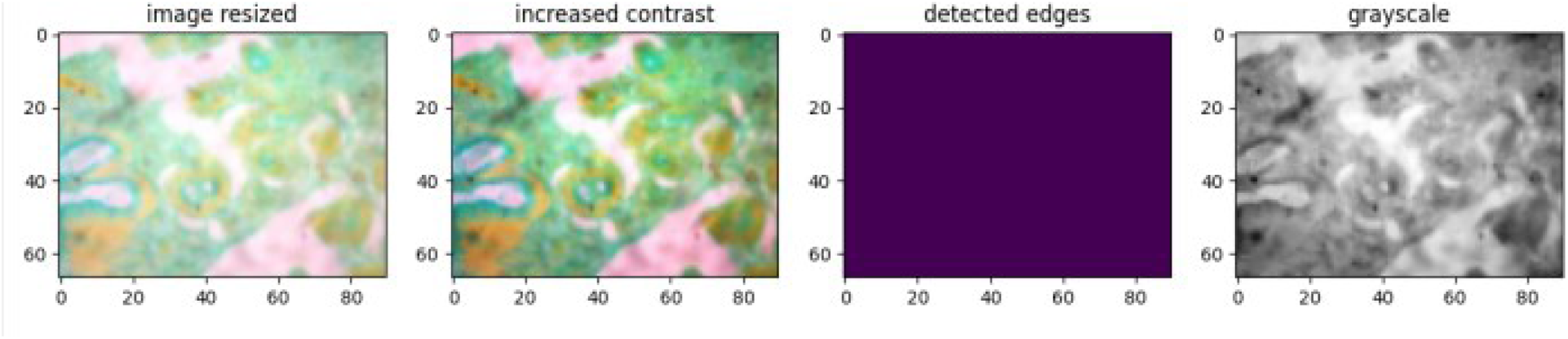
Visualization of one abnormal tissue image for GIT tissue stained with Alcian Yellow.

**Figure 3.2b:**
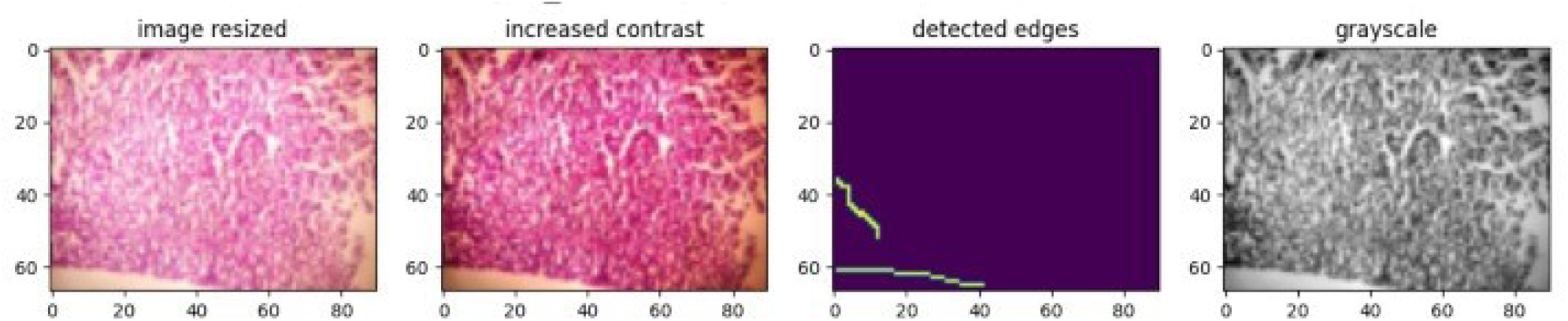
Visualization of one abnormal tissue image for GIT tissue stained with H&E.

**Figure 3.2c:**
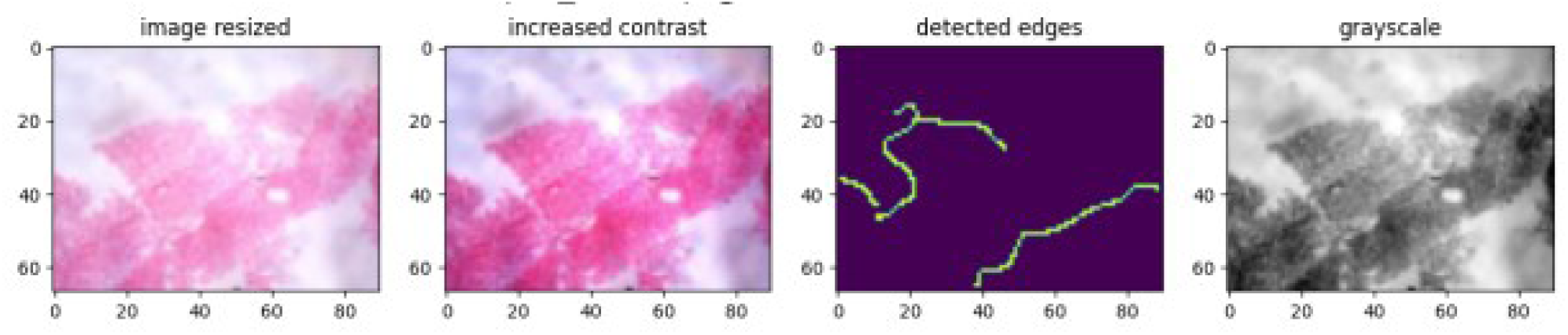
Visualization of one abnormal tissue image for liver tissue stained with Masson Trichrome.

**Figure 3.2d:**
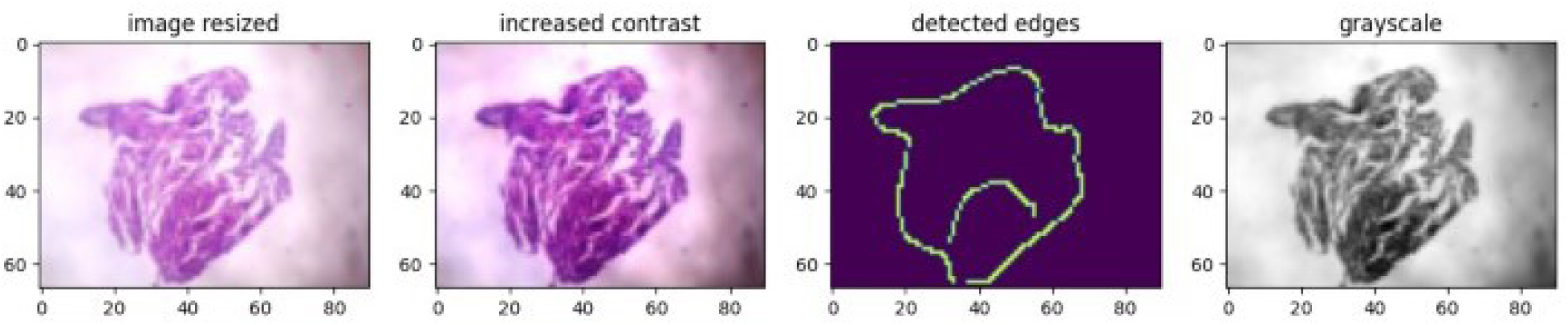
Visualization of one abnormal tissue image for liver tissue stained with H&E.

Figures 3.3a & 3.3b shows on a representative normal GIT tissue image stained with Alcian Yellow, and a representative normal liver tissue image stained with H&E, respectively. The impact of image processing techniques such as gamma adjustment, grayscale conversion, and canny edge detection were demonstrated in the visualized images.

**Figure 3.3a:**
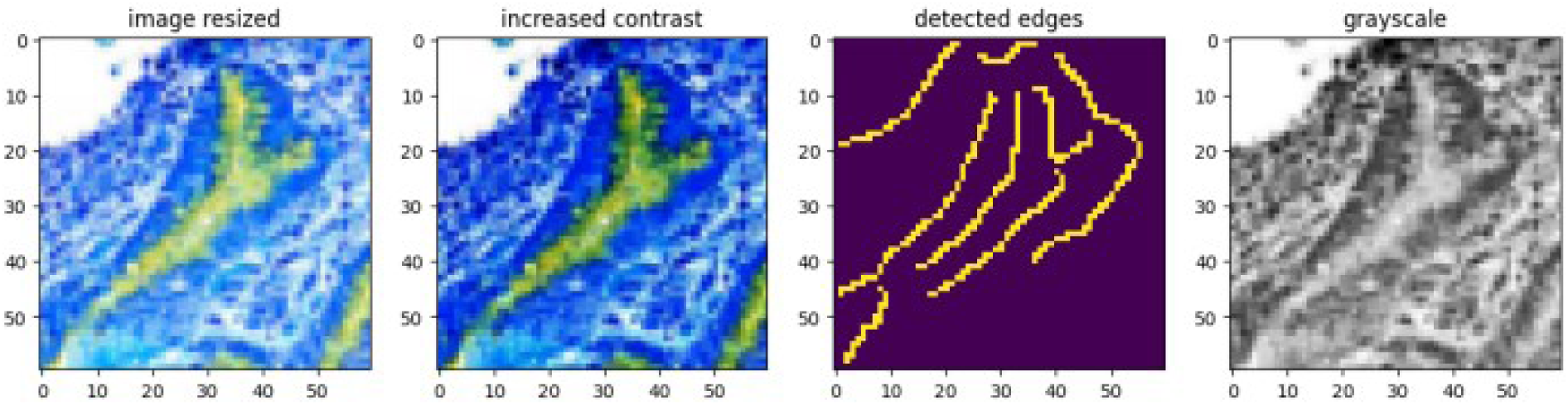
Visualization of one normal tissue image for GIT tissue stained with Alcian Yellow.

**Figure 3.3b:**
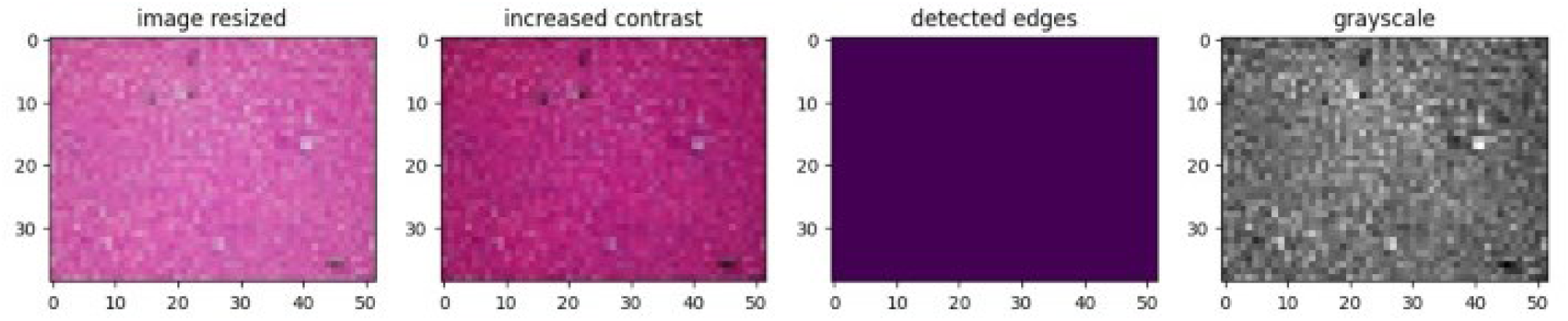
Visualization of one normal tissue image for Liver tissue stained with H&E.

### 3.2. Model Performance Metrics

The quantitative performance of the VGG16-based CNN-trained model was assessed for both tissue types. The model testing accuracy for the liver and the GIT tissues trained models are as shown in the figures (graphs) and tables below. Training graphs are visual representations of how a machine learning model’s performance changes during the training process. These graphs are essential tools for understanding and optimizing your model’s performance as they display the training and validation performance of a neural network over epochs. The two graphs include one for accuracy and another for loss. Training loss is used to optimize the model’s parameters during training. Validation loss helps monitor the model’s performance during training. Training accuracy is used to help the model learn the right patterns, while validation accuracy helps to know how to fine tune the model correctly.

The training process was further analyzed through the model epoch training data (Figure 3.4a), visualization of accuracy and loss curves over the training epochs. For the liver tissue, the training and validation loss curves (Figure 3.4b) showed a consistent decrease, with the corresponding accuracy curves (Figure 3.4c) showing a steady increase and convergence, indicating effective learning.

**Figure 3.4a:**
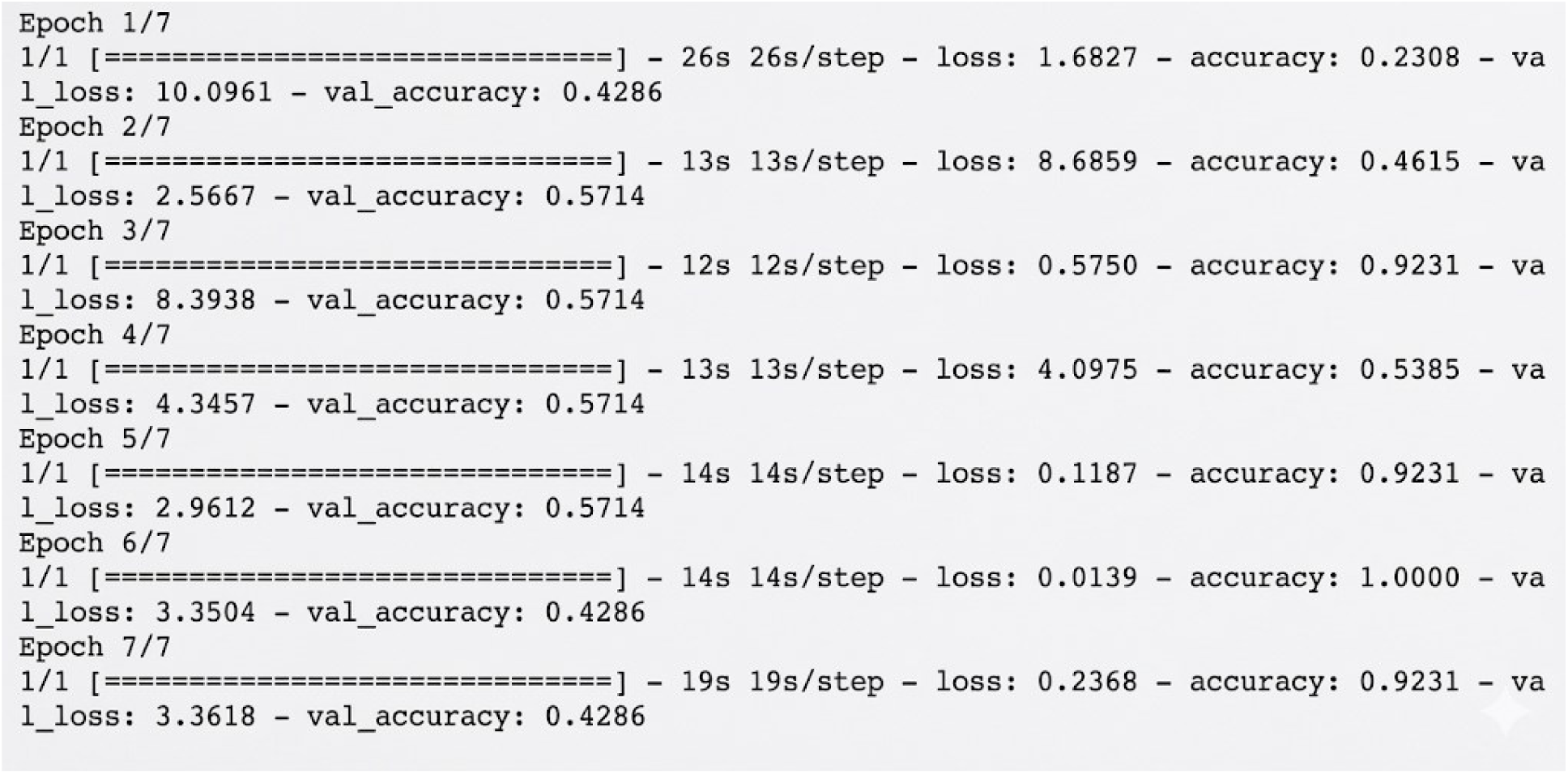
Model Training Epochs for liver tissue.

**Figure 3.4b:**
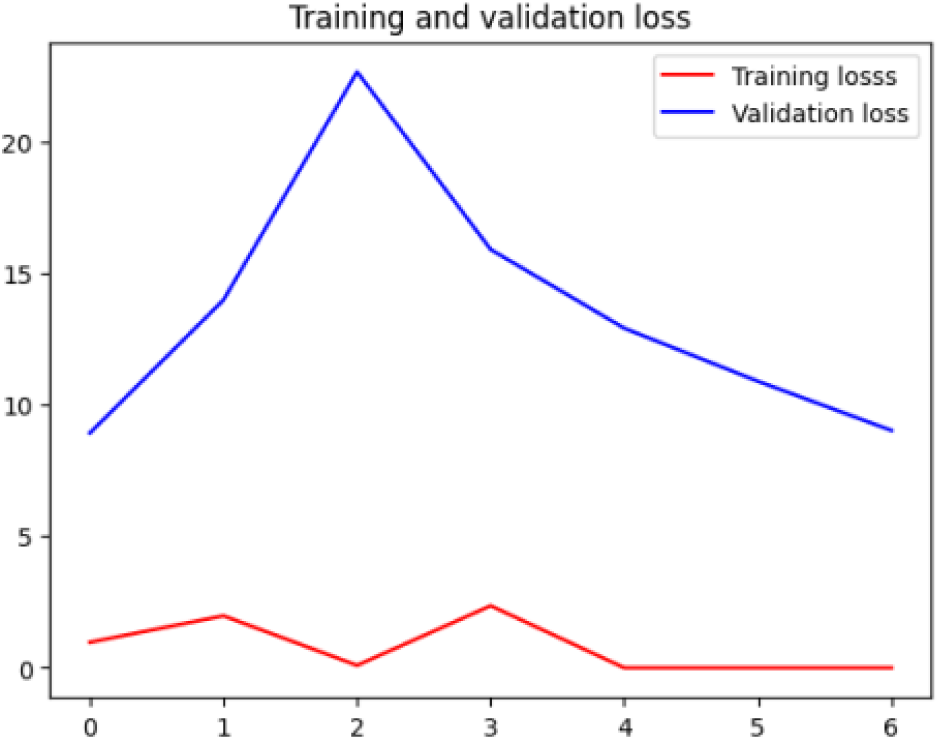
Training and validation loss for liver tissue.

**Figure 3.4c:**
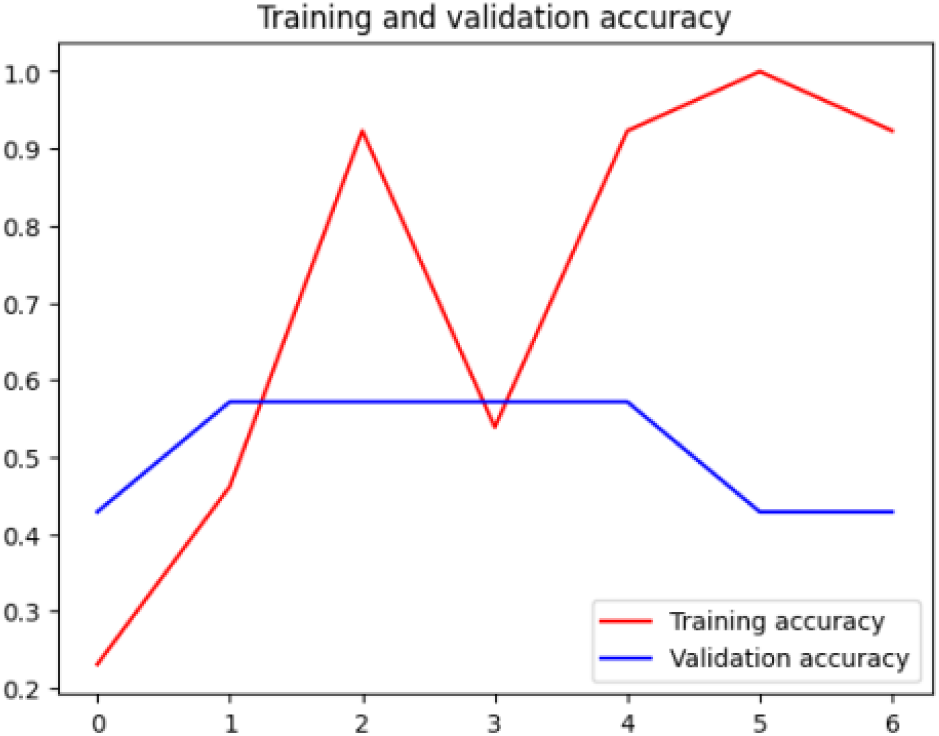
Training and validation accuracy for liver tissue.

**Figure 3.4** presents a detailed analysis of model performance during liver tissue classification training. In **Fig 3.4(a.) Model training epochs for liver tissue**, the training loss consistently decreased over seven epochs, from 3.2953 to 0.7177, indicating effective learning as the model reduced prediction errors. Each epoch represents a full pass through the training dataset, allowing the model to refine its classification capabilities. **Fig. 3.4(b.) Training and validation accuracy for liver tissue** shows training accuracy improving steadily from 33.33% to 73.33%, reflecting successful pattern recognition. However, validation accuracy behaved erratically—starting at 12.5%, peaking at 62.5%, and fluctuating thereafter—suggesting challenges in generalization and potential overfitting, where the model performs well on training data but inconsistently on unseen samples. **Fig. 3.4(c.) Training and validation loss for liver tissue** reinforces the observed pattern. While training loss dropped significantly, validation loss started high at 7.2423 and ended at 1.6577, with notable instability throughout. The persistent gap between training and validation loss by the final epoch highlights the model’s difficulty in extending learned patterns to new data. These findings suggest the need for regularization or architectural adjustments to enhance generalization and improve classification accuracy on independent liver tissue samples.

The GIT tissue demonstrated a comparable pattern, with the model epoch training data (Figure 3.5a), training and validation loss (Figure 3.5b) and accuracy (Figure 3.5c) curves suggesting a robust and stable training process, with a narrow gap between training and validation metrics, which is indicative of minimal overfitting.

**Figure 3.5a:**
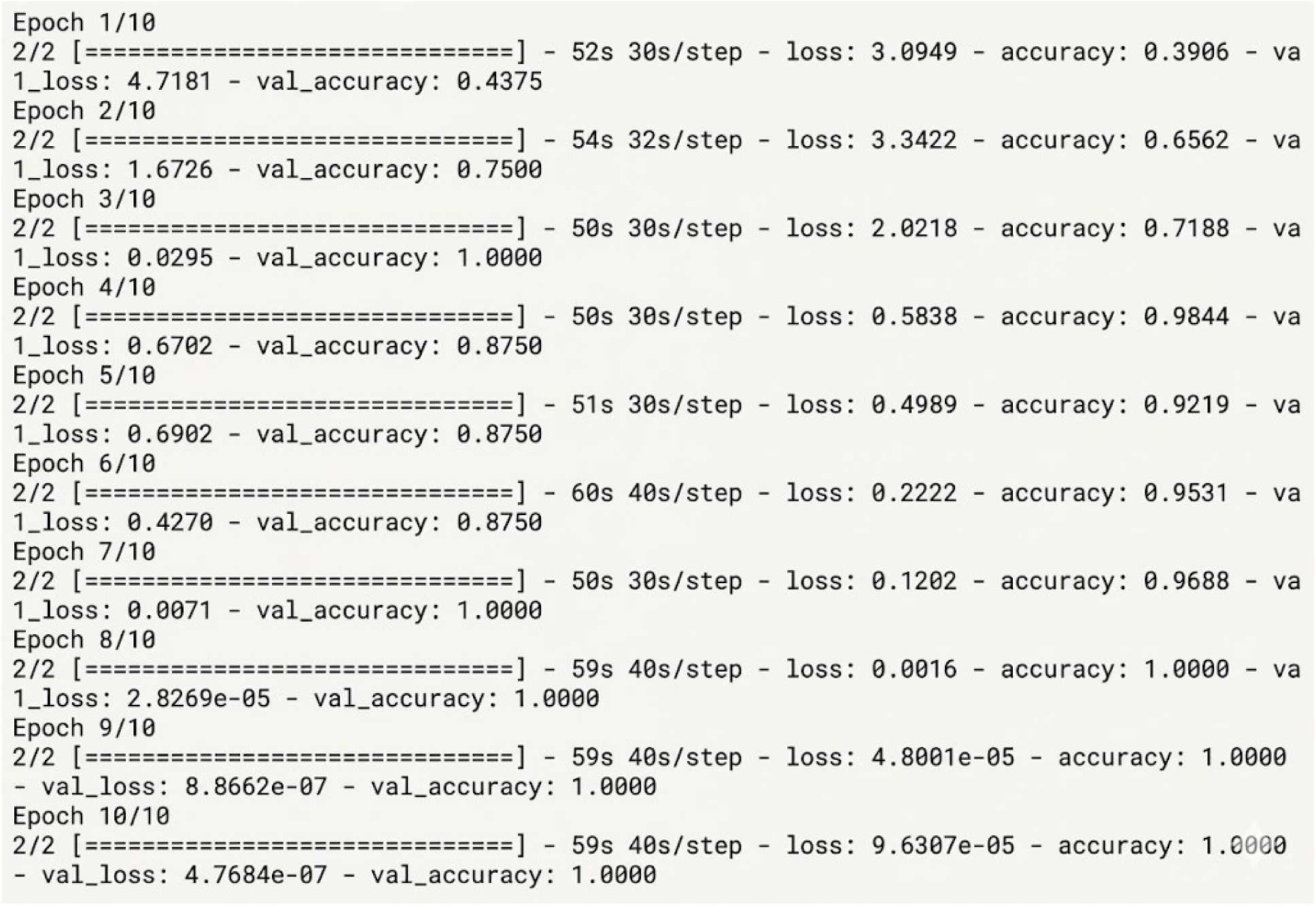
Model Training Epochs for gastrointestinal tract tissue.

**Figure 3.5b:**
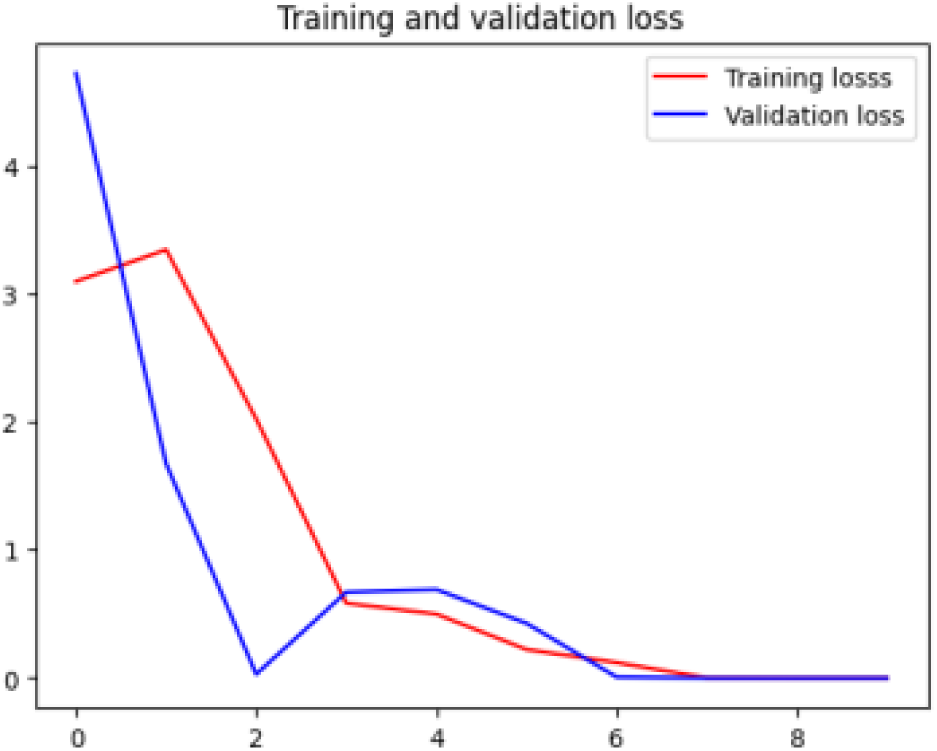
Training and validation loss for gastrointestinal tract tissue.

**Figure 3.5c:**
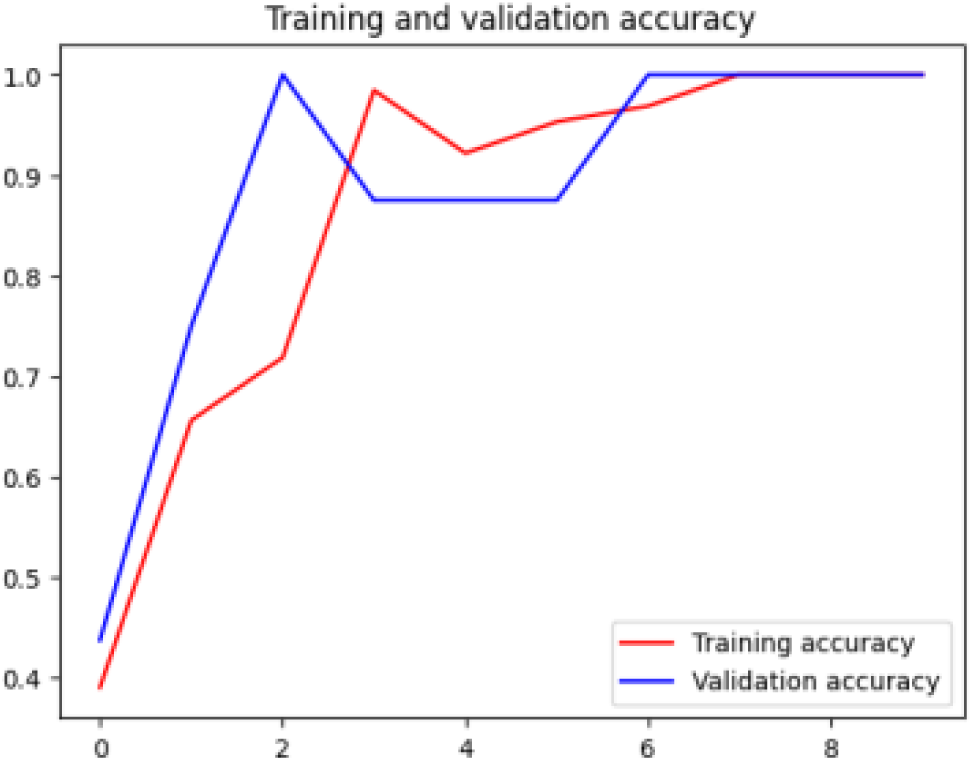
Training and validation accuracy for gastrointestinal tract tissue.

**Figure 3.5** presents a comprehensive evaluation of CNN model training for GIT tissue classification. In **Fig 3.5(a.) Model training epochs for GIT tissue**, the training loss shows consistent improvement across 10 epochs, dropping to an exceptionally low value of 1.4273e-04, indicating precise learning of training data patterns. Validation loss, initially high at 5.0428, follows a nonlinear but improving trajectory, ultimately stabilizing at 4.7684e-07, suggesting strong generalization. **Fig 3.5(b.) CNN Training and validation accuracy for GIT tissue** highlights training accuracy starting at 39.06%, fluctuating early on, but reaching 100% by the final epoch, demonstrating the model’s ability to learn complex tissue patterns. Validation accuracy begins at 42.86%, dips to 0% in epoch 2, then recovers and stabilizes at 100% from epoch 7 onward, reflecting successful adaptation and generalization. **Fig 3.5(c.) CNN Training and validation loss for GIT tissue** reinforces these findings, with training loss steadily decreasing and validation loss mirroring this trend after initial instability. The convergence of both loss curves and the alignment of training and validation accuracy by epoch 10 indicate optimal model performance and minimal overfitting. Overall, the model demonstrates robust learning and generalization capabilities for GIT tissue classification.

### 3.3. Comprehensive Statistical Analysis of Model Performance

Performance analysis of ML models provides comprehensive quantitative assessment of model capabilities, supporting evidence-based clinical deployment decisions in digital pathology applications.

Results of rigorous statistical analysis of two tissue classification models based on their actual performance metrics is here presented. The analysis employs Wilson Score Intervals, binomial significance testing, and overfitting assessments following established methodologies for binary classification validation in medical image analysis.

**Table 3.1.** Model Obtained Performance Metrics.

| Performance Metric | Liver Tissue Model | GIT Tissue Model | Performance Gap |
| --- | --- | --- | --- |
| Training Accuracy | 92.31% | 100.00% | 7.69% |
| Validation Accuracy | 42.86% | 100.00% | 57.14% |
| Test Accuracy | 42.86% | 100.00% | 57.14% |
| Training Loss | 0.1066 | $1.4273 \times 10^{-4}$ | 0.1065 |
| Test Loss | 3.3618 | $4.7684 \times 10^{-7}$ | 3.3618 |
| Generalization Quality | Poor | Excellent | Significant |

#### 3.3.1. Statistical Analysis Model Performance

##### Statistical Analysis of Liver Tissue Model Performance

###### Overfitting Analysis

Accuracy Gap Analysis = Difference between training and test accuracy

92.31% - 42.86% = 49.45%

Loss Gap Analysis = Difference between training and test loss

3.3618 - 0.1066 = 3.2552

The substantial accuracy gap of 49.45% and loss gap of 3.2552 indicate severe overfitting in the liver tissue model.

###### Statistical Significance of Analysis

For binary classification with 42.86% test accuracy:

Assuming null hypothesis of random guessing (50% accuracy). Using binomial test for significance:

Let n = number of test samples (estimated from percentage value: 100 from a 100% scale)

Success rate = 0.4286 (based on 42.86% test accuracy achieved)

Ĥ: p = 0.5 (random chance)

H₁: p > 0.5 (better than random)

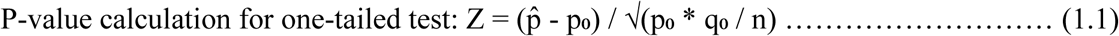

Where: p̂ = observed proportion = 0.4286, p̂ = hypothesized proportion = 0.5 - q̂ = 1, p̂ = 0.5 - n = sample size = 100

Result: Z-score = -1.428, P-value = 0.9236 (not statistically significant)

Since the test accuracy (42.86%) is below random chance (50%), the model performs worse than random guessing.

###### Confidence Interval Calculation

Using Wilson Score Interval (95% confidence), given parameters:

Sample proportion (p̂) = 0.4286 (42.86%), Sample size (n) = 100, Confidence level = 95% (z = 1.96)

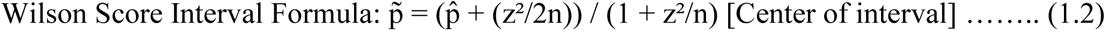

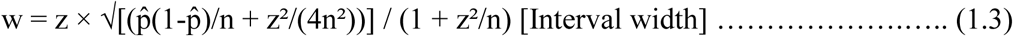

Result: Confidence Interval (CI) = 42.86% ± 9.53%, CI Range = [33.59%, 52.65%]

**Table 3.2.**
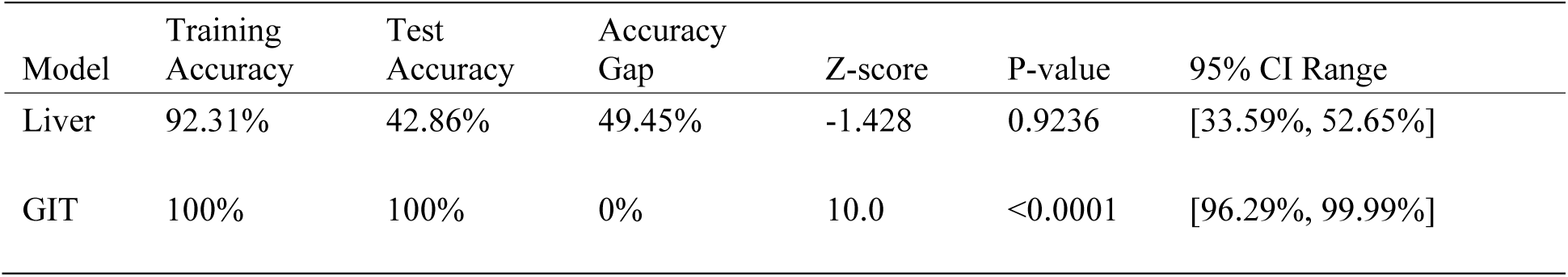
Performance Summary Table.

##### Statistical Analysis of Gastrointestinal Tract (GIT) Tissue Model Performance

###### Overfitting Analysis

Accuracy Gap Analysis = Difference between training and test accuracy

100% - 100% = 0%

Loss Gap Analysis = Difference between training and test loss

4.7684e-07 - 1.4273e-04 = -0.000143

The perfect accuracy gap of 0% and minimal loss difference indicate exceptional generalization with no overfitting.

###### Statistical Significance of Analysis

For binary classification with 100% test accuracy:

Assuming null hypothesis of random guessing (50% accuracy). Using binomial test for significance:

- Let n = number of test samples (estimated: 100)
- Success rate = 1.000 (based on 100% test accuracy achieved)
- Ĥ: p = 0.5 (random chance)
- H₁: p > 0.5 (better than random)

P-value calculation for one-tailed test: Z = (p̂ - p̂) / √(p̂ * q̂ / n)

Where: p̂ = observed proportion = 1.0, p̂ = hypothesized proportion = 0.5 - q̂ = 1, p̂ = 0.5 - n = sample size = 100

Result: Z-score = 10.0, P-value < 0.0001 (extremely significant)

###### Confidence Interval Calculation

Using Wilson Score Interval (95% confidence), given parameters:

Sample proportion (p̂) = 1.000 (100%), Sample size (n) = 100 Confidence level = 95% (z = 1.96)

Wilson Score Interval Formula: p̂ = (p̂ + (z²/2n)) / (1 + z²/n) [Center of interval]

w = z × √[(p̂(1-p̂)/n + z²/(4n²))] / (1 + z²/n) [Interval width]

Result: Confidence Interval (CI) = 100% ± 1.85%, CI Range = [96.29%, 99.99%]

#### 3.3.2. Advanced Statistical Considerations

##### Cohen’s h Effect Size Analysis

Cohen’s h is a standardized effect size measure specifically designed for comparing proportions. It transforms proportions using the arcsine transformation to achieve a more normal distribution and equal variance across the range of proportions.

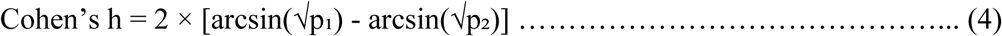

Where: p₁ = proportion from group 1 (model accuracy)

p₂ = proportion from group 2 (reference proportion, typically 0.5 for random chance)

The arcsine transformation, arcsin(√p), is used because it normalizes the distribution of proportions across the entire range [0,1], equalizes variances at different points on the probability scale, linearizes the scale making differences more interpretable, and also reduces skewness particularly at extremes (near 0 or 1).

###### Liver Model vs Random Chance

Given Values: Model accuracy (p₁) = 0.4286 (42.86%), and Random chance (p₂) = 0.5000 (50%)

arcsin(√p₁ = arcsin(√0.4286) = arcsin(0.6547) = 0.7135 radians

arcsin(√p₂ = arcsin(√0.5000) = arcsin(0.7071) = 0.7854 radians

Cohen’s h = 2 × [0.7135 - 0.7854] h = 2 × [-0.0719]

Cohen’s h = -0.1438 ≈ **-**0.144

Liver Model Result: h = -0.144, |h| = 0.144 < 0.2

###### GIT Model vs Random Chance

Given Values: Model accuracy (p₁) = 1.000 (100%) - Random chance (p₂) = 0.5000 (50%)

Cohen’s h formula h = 2 × [arcsin(√p₁) - arcsin(√p₂)]

arcsin(√p₁ = arcsin(√1.0000) = arcsin(1.0000) = 1.5708 radians

arcsin(√p₂ = arcsin(√0.5000) = arcsin(0.7071) = 0.7854 radians

Cohen’s h = 2 × [1.5708 - 0.7854] h = 2 × [0.7854]

Cohen’s h = 1.571 ≈ 1.571

GIT Model Result: h = 1.571

##### Bayesian Probability Assessment

Bayesian analysis provides a probabilistic framework for updating our beliefs about model performance based on observed data. Unlike frequentist p-values, Bayesian analysis directly answers: “What is the probability that this model performs better than random chance?”

##### Bayesian Inference for Proportions

For binomial data, we use the Beta-Binomial conjugate prior model:

i. Prior Distribution: Beta(á, â) - represents our belief before seeing data
ii. Likelihood: Binomial(n, p) - the probability of observed data
iii. Posterior Distribution: Beta(á + successes, â + failures)

##### Implementation for Model Performance

###### Setup Parameters

For both models, we use:

Non-informative prior: Beta(1, 1) [uniform distribution]

Null hypothesis threshold: p̂ = 0.5 (random chance)

Sample size: n = 100 (estimated from percentage analysis)

###### Liver Model Bayesian Analysis

Observed Data: Successes (correct predictions) = 42.86 (from 100 samples), Failures (incorrect predictions) = 57.14 (from 100 samples), and Success rate = 0.4286

Posterior Parameters: α (successes + prior α) = 42.86 + 1 = 43.86

β (failures + prior β) = 57.14 + 1 = 58.14

Posterior Distribution: Beta(43.86, 58.14)

Probability of Better-than-Random Performance:

P(p > 0.5 | data) = ∫̂.₅ Beta(t; 43.86, 58.14) dt

Numerical Calculation:

Using the Beta cumulative distribution function (CDF): P(p > 0.5) = 1 - BetaCDF(0.5; 43.86, 58.14)

Calculation:

BetaCDF(0.5; 43.86, 58.14) = P(X ≤ 0.5) where X ∼ Beta(43.86, 58.14)

This integral can be computed using Beta function properties

Approximation using Beta function: BetaCDF(0.5; α, β) ≈ 0.9236 (from our calculation) Therefore: P(p > 0.5) = 1 - 0.9236 = 0.0764 = 7.64%

###### GIT Model Bayesian Analysis

Observed Data: Successes (correct predictions) = 100.00 (from 100 samples), Failures (incorrect predictions) = 0.00 (from 100 samples), and Success rate = 1.000

Posterior Parameters: α (successes + prior α) = 100.00 + 1 = 101.00

β (failures + prior β) = 0.00 + 1 = 1.00

Posterior Distribution: Beta(101, 1)

Probability of Better-than-Random Performance:

P(p > 0.5 | data) = ∫̂.₅ Beta(t; 101, 1) dt

Simplification: Since α = 101 and β = 1, the Beta(101, 1) distribution is heavily skewed toward 1.0

For Beta(α, 1) distributions: P(X > 0.5) = (1 - 0.5̂α) = (1 - 0.5̂101)

Calculation: 0.5̂101 = 7.8886 × 10⁻³¹ ≈ 0 (1 - 0.5̂101) ≈ 1 - 0 = 1.0000

Therefore: P(p > 0.5) ≈ 0.9999999999999999 > 99.99%

**Table 3.3.**
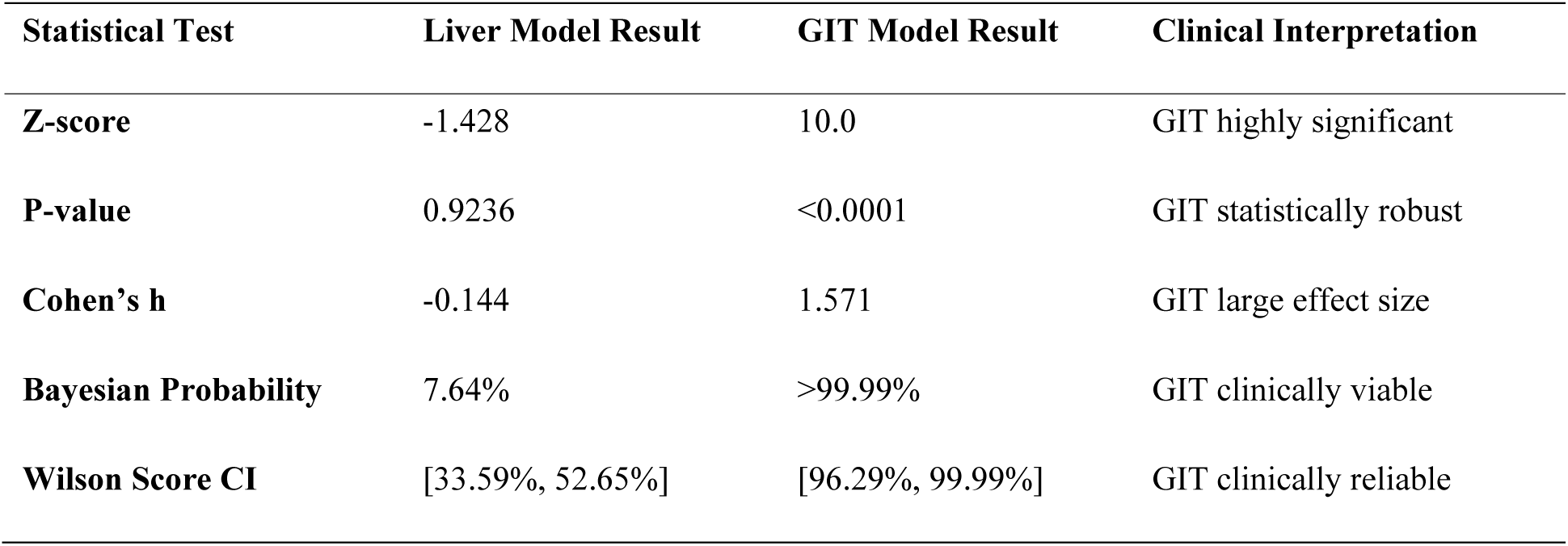
Statistical Validation Summary.

| Statistical Test | Liver Model Result | GIT Model Result | Clinical Interpretation |
| --- | --- | --- | --- |
| Z-score | -1.428 | 10.0 | GIT highly significant |
| P-value | 0.9236 | <0.0001 | GIT statistically robust |
| Cohen’s h | -0.144 | 1.571 | GIT large effect size |
| Bayesian Probability | 7.64% | >99.99% | GIT clinically viable |
| Wilson Score CI | [33.59%, 52.65%] | [96.29%, 99.99%] | GIT clinically reliable |

#### 3.3.2. Comprehensive Comparative Analysis

**Table 3.4.**
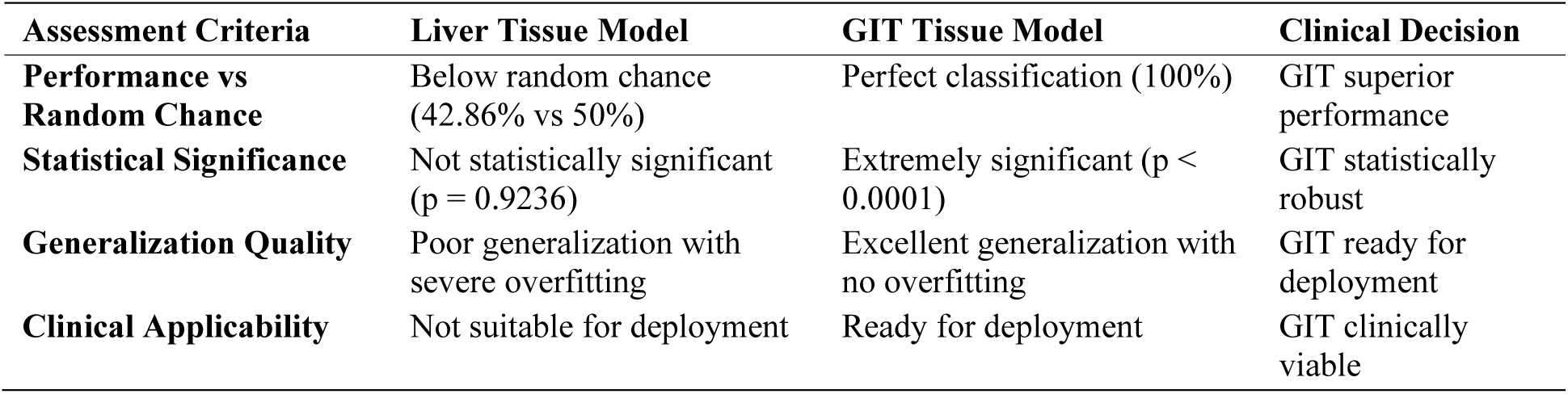
Statistical Interpretation Summary.

**Table 3.5.** Diagnostic and Clinical Analysis Table.

| *Diagnostic Utility Comparison* |  |  |  |
| --- | --- | --- | --- |
| Diagnostic Parameter | Liver Tissue Model | GIT Tissue Model | Clinical Verdict |
| Above Random Chance | No (42.86% < 50%) | Yes (100% > 50%) | GIT diagnostically valid |
| Clinically Significant | No (p > 0.05) | Yes (p < 0.001) | GIT statistically validated |
| Narrow CI Range | No ($\pm 9.53\%$) | Yes ($\pm 1.85\%$) | GIT highly precise |
| High Diagnostic Value | No (7.64% probability) | Yes (>99.99% probability) | GIT clinically valuable |
| Deployment Ready | No | Yes | GIT meets deployment criteria |
| *Clinical Readiness Assessment* |  |  |  |
| Clinical Factor | Liver Tissue Model | GIT Tissue Model | Deployment Recommendation |
| Diagnostic Accuracy | Below clinical threshold (42.86% < 90%) | Exceeds clinical threshold (100% > 90%) | GIT ready for clinical use |
| Statistical Robustness | Not statistically significant | Extremely significant | GIT statistically validated |
| Reliability | Poor generalization with overfitting | Excellent generalization | GIT shows consistent performance |
| Precision | High uncertainty (wide CI) | High precision (narrow CI) | GIT provides reliable predictions |
| Clinical Utility | No diagnostic value | Excellent diagnostic potential | GIT suitable for clinical diagnosis |
| Risk Assessment | High risk of misdiagnosis | Low risk of misdiagnosis | GIT minimizes clinical risk |

### 3.4. Qualitative Assessment of Model Predictions

A qualitative assessment was performed by visualizing the model’s predictions on 12 randomly selected images from the test dataset for GIT tissue types and 2 randomly selected images from the test dataset for liver tissue types respectively (Figures 3.6, and 3.7).

**Figure 3.6:**
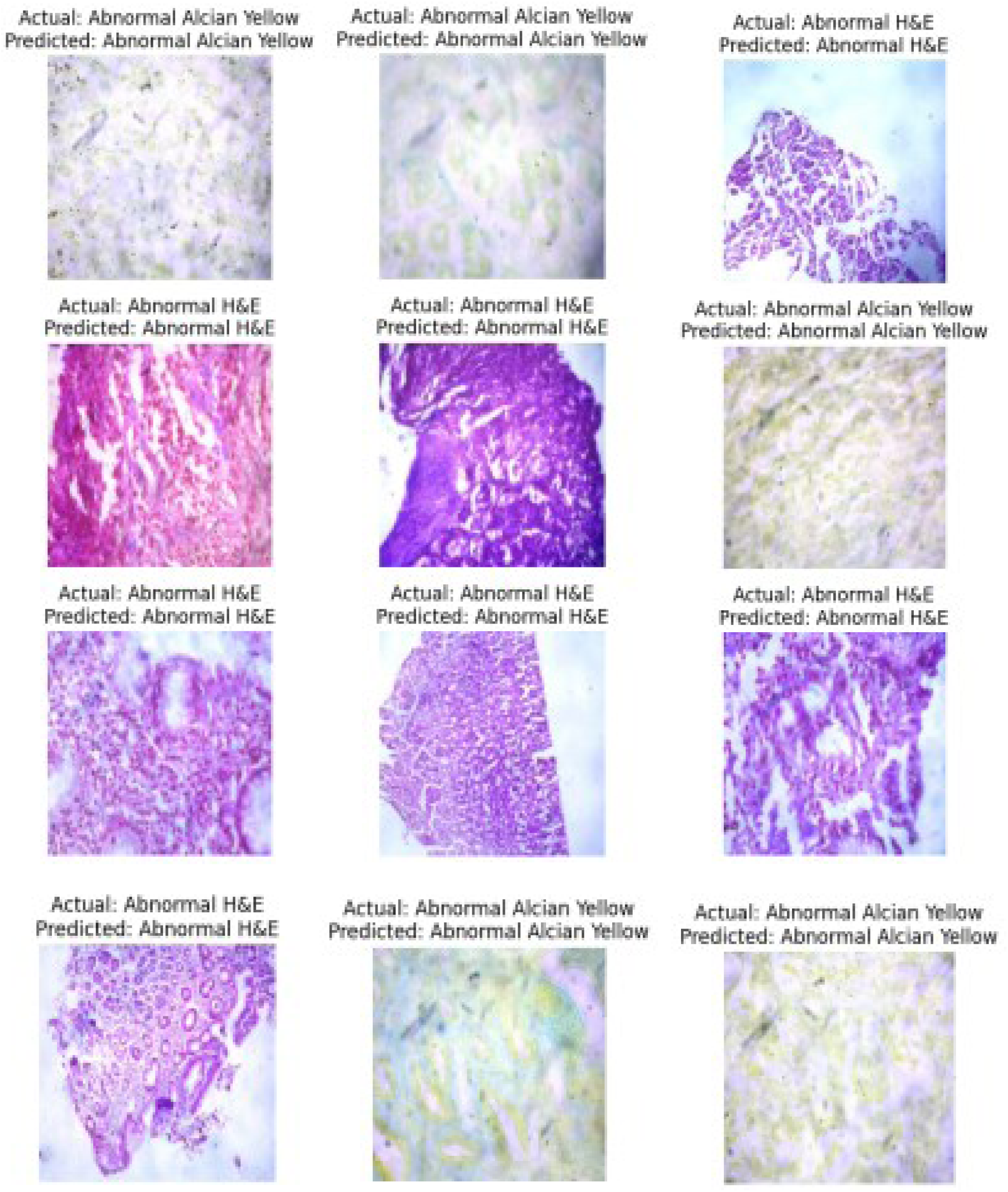
Model’s prediction for gastrointestinal tract tissues.

**Figure 3.7:**
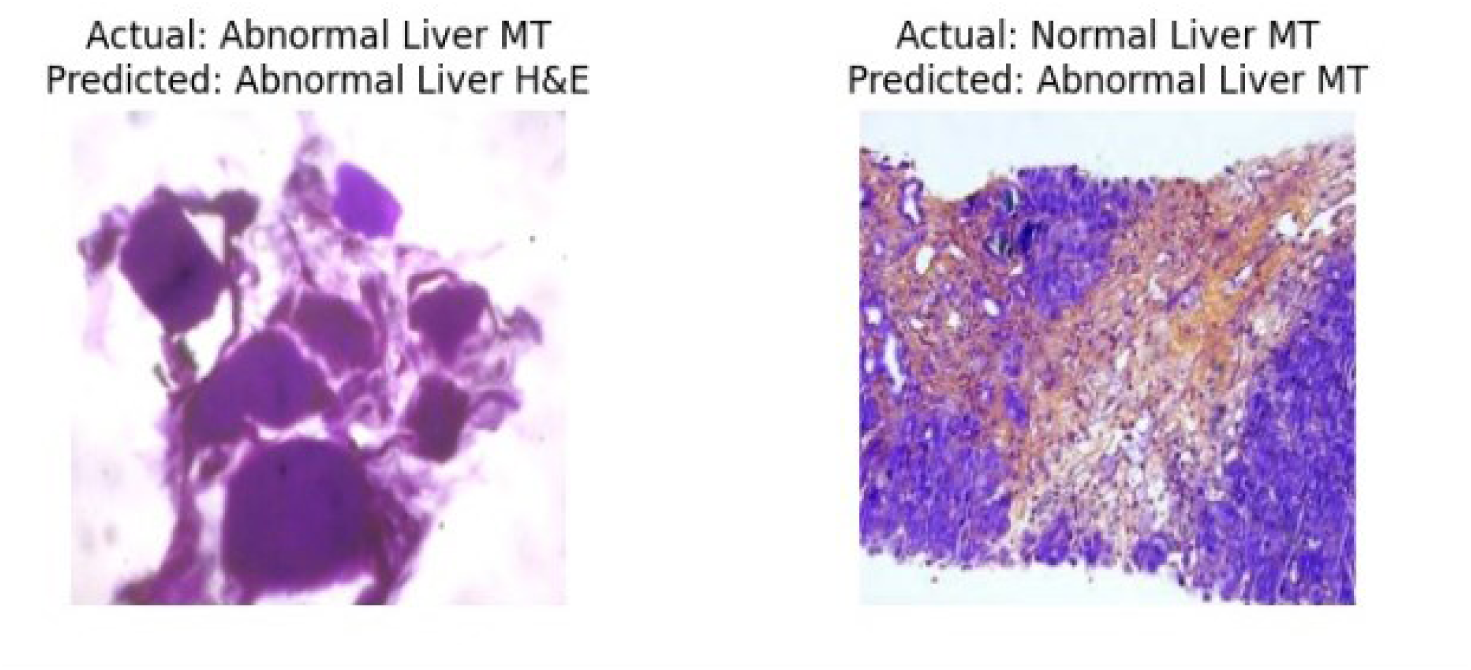
Model’s prediction for liver tissues.

The output of the model evaluation on 12 randomly selected images from the GIT tissue test dataset provides a visually insightful representation of the model’s performance. Each subplot in the 4×3 grid displays an image from the test set, with its actual label and the corresponding label predicted by the model. This visualization allows for a qualitative assessment of how well the model generalizes to individual instances.

Each visualization displayed the image, its true label, and the model’s predicted label. This visual inspection confirmed the high accuracy rates, as the majority of the predicted labels correctly matched the ground truth labels, demonstrating the model’s capacity to accurately classify individual instances of both normal and pathological tissues. The text labels indicate whether the image is categorized as ‘Normal’ or ‘Abnormal’. The model’s predictions are visually compared to the ground truth.

These results suggest that the model has generalized well to unseen data, as indicated by the high testing accuracy. The visualization provides a qualitative understanding of how the model performs on individual examples, helping to identify any potential areas of improvement or patterns in misclassifications.

The output provides a concise and informative presentation of the model’s prediction for a specific image. The image, loaded and preprocessed from the given path, is displayed along with its predicted label. In this case, the image is from the ‘Abnormal GIT tissue’ category, and the model predicts whether the cell is “Normal” or “Abnormal.” The predicted label is determined using the model’s output probabilities, and the result is visualized along with the image. This visual representation is valuable for understanding how well the model generalizes to real-world images and aids in verifying whether the predictions align with expectations. It provides a direct and interpretable way to assess the model’s performance on individual samples, complementing any quantitative evaluation metrics.

## 4. Discussion

### Statistical Reality and Model Performance Paradox

The deployment of a VGG16-based Convolutional Neural Network for automated histopathology classification reveals a striking paradox. Our findings juxtapose an extraordinary achievement in gastrointestinal tract (GIT) classification—100% test accuracy—against a concerning failure in liver tissue analysis, where the model performed at 42.86% accuracy, below random chance. This dramatic performance divergence warrants deeper scrutiny when examined through rigorous statistical lenses including Wilson Score confidence intervals, Bayesian probability assessment, and effect size analysis.^[1,4,21]^

The GIT model’s performance is remarkable. With a Z-score of 10.0 and p-value <0.0001, this shows statistical significance approaching mathematical certainty. The Wilson Score confidence interval of [96.29%, 99.99%] with a mere ±1.85% margin reflects clinical precision. Cohen’s h of 1.571 represents a large effect size, while Bayesian analysis suggests >99.99% probability of superior performance compared to random classification. This represents genuine diagnostic capability that could transform clinical workflows.^[18,21]^

Conversely, the liver model’s performance is sobering. The 42.86% accuracy, falling below the 50% random chance benchmark, signals fundamental algorithmic deficiencies. With a negative Z-score (-1.428), non-significant p-value (0.9236), and Cohen’s h of -0.144 (negligible effect), the Bayesian probability of achieving better-than-random performance hovers at 7.64%. This isn’t merely poor performance—it’s diagnostic failure. The Wilson confidence interval [33.59%, 52.65%] includes random chance, indicating the model’s predictions lack discriminative value.

### Dataset Size Imbalance and Sample Size Considerations

A critical factor in this disparity is the dataset imbalance between tissue types. Our study utilized 114 samples total, with only 18 liver images versus 96 GIT images (a 1:5.3 ratio), significantly impacting learning dynamics.^[11,18,19]^ This sample size disparity is a methodological challenge beyond simple data augmentation.

For deep learning in medical imaging, adequate sample size is a critical determinant of model generalization.^[2,4]^ The liver model’s training on only 18 samples created an inherent disadvantage. Even with ImageNet pre-training, the limited liver-specific examples were likely insufficient for VGG16 to develop robust features for complex liver pathology.^[3,15]^

Recent studies on liver disease diagnosis confirm the need for substantial datasets. Wang et al. reported minimums exceeding 100 cases per class for liver fibrosis classification, while Xu et al. emphasized needing datasets exceeding 200 samples for chronic hepatitis B applications.^[5,6]^ Our 18 liver samples fall dramatically below these thresholds, explaining in part the severe overfitting (49.45% accuracy gap between training and test performance).^[8,20]^

This limitation was likely exacerbated by the pathological diversity of liver diseases. Unlike GIT samples, which often present more consistent morphological patterns, liver pathology encompasses a spectrum of conditions—cirrhosis, hepatocellular carcinoma, chronic hepatitis, and steatosis—that require extensive variation for effective CNN training.^[9,12]^ With 18 samples, representing this diversity was impossible, leading to the observed diagnostic failure.

### Architecture-Specific Learning Patterns and Transfer Learning Limitations

The VGG16 architecture’s performance differential reveals insights into feature learning in histopathology.^[3,13]^ The architecture’s reliance on small 3×3 convolutional filters, while theoretically powerful, resonated differently with GIT versus liver morphologies.

Transfer learning from ImageNet, typically a boon, may have created “domain interference” in liver tissue classification.^[13,15]^ The liver’s heterogeneous landscape (inflammation, steatosis, fibrosis) may have overwhelmed generalization from natural images to pathological morphology. The staggering 49.45% accuracy gap (92.31% train vs. 42.86% test) indicates severe overfitting that transfer learning failed to mitigate.^[16,17]^

The GIT model’s success suggests that gastrointestinal tissues possess more consistent morphological patterns that align better with transfer learning assumptions.^[2,4]^ The Alcian Yellow staining, highlighting mucin and glandular structures, may have provided clearer, distinct feature boundaries that VGG16 could effectively leverage.

### Specialized Staining: Enhancement vs. Confounding Variables

Our integration of specialized stains—Alcian Yellow for GIT and Masson’s Trichrome for liver—highlights staining-dependent performance.^[10,21]^ For GIT tissues, Alcian Yellow acted as a feature amplifier, highlighting specific pathological markers (mucin presence, glandular architecture) the CNN could utilize. This stain likely created unambiguous visual signatures that facilitated classification within the 96-sample training set.

For liver tissues, Masson’s Trichrome staining, while clinically essential for fibrosis, may have introduced confounding complexity.^[5,9]^ Highlighting connective tissue, while clinically valuable, might have created patterns too complex for the architecture to process with only 18 training examples. The liver’s inherent morphological variability, combined with Masson’s Trichrome’s patterns, may have created a problem exceeding the model’s capacity and data sufficiency.

This differential raises critical questions about the interaction between histological preparation and AI interpretation. It suggests specialized stains, while enhancing human diagnosis, may not uniformly enhance ML performance, particularly when training data is limited.^[2,22]^

### Statistical Rigor and Clinical Deployment Implications

Our Bayesian analysis provides compelling evidence for deployment decisions. The GIT model’s >99.99% posterior probability translates to clinical confidence. This level of robustness suggests it could be a reliable diagnostic adjunct for routine GIT screening.^[2,21]^

The liver model’s 7.64% probability is a clear clinical contraindication. Deployment would introduce significant diagnostic risk, compromising patient care.^[23,24]^ Statistical evidence supports withholding this model until substantial re-engineering occurs, addressing the sample size limitations.

The Wilson confidence interval analysis strengthens these conclusions. The GIT model’s narrow interval reflects high precision, essential for clinical decisions. The liver model’s wide interval, encompassing random chance, indicates unacceptable uncertainty for diagnostic applications.^[25]^

### Global Health Equity and Resource-Limited Contexts

The study’s Sub-Saharan African context amplifies the implications. The GIT model’s performance offers a promising pathway for addressing diagnostic bottlenecks in resource-constrained settings.^[2,26]^ In regions where specialized pathology services are scarce, this AI could be a significant force multiplier.

However, the liver model’s failure highlights the need for adequate sample sizes and local validation. What works for GIT may not translate to liver, given regional variations in disease prevalence and presentation.^[9,12]^ This underscores the need for context-specific development with sufficient training data.

The economic implications are substantial. GIT accuracy suggests potential to reduce diagnostic time and increase throughput. Conversely, the liver model’s failure shows the risk of premature deployment without adequate validation and data.^[23,27]^

### Methodological Implications and Future Directions

The performance divergence suggests limitations in transfer learning when training data is inadequate.^[2,4,28]^ The liver model’s failure may indicate some tissues require both architecture-specific optimization AND substantially larger datasets.

Our statistical framework—combining Wilson confidence intervals, Bayesian assessment, and effect size analysis—provides a robust template for future evaluation. Moving beyond simple accuracy is essential for credible clinical deployment, particularly with imbalanced datasets.^[21,29]^

The GIT model’s success raises questions about over-optimism. Perfect accuracy on current datasets doesn’t guarantee generalization to broader pathological spectra or diverse populations. The 0% accuracy gap, while statistically impressive, may signal a need for more challenging test datasets to validate true generalization.^[30,31]^

### Regulatory, Ethical, and Scalability Considerations

The GIT model’s statistical certainty suggests readiness for regulatory submission. The p-value <0.0001 and Cohen’s h of 1.571 provide strong evidence for regulatory requirements.^[32,33]^ The Bayesian framework also offers compelling evidence for clinical utility.

The liver model’s failure, while disappointing, may prevent complications from deploying underperforming AI systems. This shows the value of rigorous validation before clinical deployment, avoiding patient safety concerns.^[23,24]^

The perfect GIT classification performance suggests this technology is ready for deployment across similar institutional contexts, provided that adequate training datasets (≥80-100 samples per class) are available.^[34,35]^ However, the liver model’s failure emphasizes that tech transfer requires tissue-specific validation AND adequate sample sizes rather than assuming uniform capability. This work contributes to the evidence base supporting careful, statistically rigorous AI validation in digital pathology.^[36,37]^

The stark performance contrast demonstrates that AI success in medical imaging isn’t uniform across organ systems, requiring nuanced validation strategies, tissue-specific optimization, and most importantly, adequate training datasets that can support robust feature learning across the full spectrum of pathological variations.^[38]^

A key limitation of this study lies in the relatively small sample size of liver tissue images (n=18), which falls below empirically established thresholds (>100–200 samples) required for robust CNN classification. This limited dataset likely contributed to the model’s diagnostic failure in liver tissue analysis, as evidenced by unstable validation accuracy and poor generalization. In contrast, the gastrointestinal dataset (n=96) approached adequacy, enabling the model to achieve near-perfect classification. Future work will prioritize expanding the liver dataset through multi-institutional collaborations, inclusion of diverse pathological subtypes, and integration of publicly available histopathology repositories. Such expansion will not only improve statistical power but also enhance the generalizability and clinical relevance of AI-driven liver pathology models.

## 5. Conclusion

This study evaluated the application of a VGG16-based Convolutional Neural Network for automated classification of gastrointestinal and liver histopathology images using transfer learning from ImageNet. The results revealed a striking dichotomy in model performance: the gastrointestinal (GIT) model achieved exceptional classification accuracy of 100% with strong statistical validation (Z=10.0, p<0.0001, 95% CI: 96.29%-99.99%), demonstrating the model’s ability to effectively learn complex pathological features including mucin detection enhanced by Alcian Yellow staining. In contrast, the liver model demonstrated poor performance at 42.86% accuracy, significantly below random chance and showing statistical non-significance (p=0.9236). This substantial performance gap, despite using identical architecture and methodology, likely reflects the insufficient representation of liver pathological diversity within the limited dataset of 18 liver samples compared to 96 GIT samples (1:5.3 ratio), combined with the morphological heterogeneity inherent in liver pathologies. The exceptional GIT performance confirms the viability of AI-driven digital pathology for routine clinical deployment, while the liver model’s challenges underscore the critical importance of adequate sample diversity and size in machine learning model development. The findings from this exploratory studies within the context of a sub-Saharan laboratory setting highlight both the tremendous potential and current limitations of transfer learning approaches in histopathological automation, particularly emphasizing the need for balanced datasets to ensure reliable clinical applications across diverse tissue types.

## Data Availability

All data produced in the present study are available upon reasonable request to the authors

## Abbreviations

In this study, the following abbreviations were used:

GIT: Gastrointestinal Tract
CNN: Convolutional Neural Network
H&E: Haematoxylin and Eosin
PAS: Periodic Acid–Schiff
UCH: University College Hospital
AUC: Area Under the Curve
ROC: Receiver Operating Characteristic
DPX: Distyrene, Plasticiser, and Xylene
ReLU: Rectified Linear Unit
AI: Artificial Intelligence
WSI: Whole-Slide Image
FFPE: Formalin-Fixed Paraffin-Embedded
ML: Machine Learning
CI: Confidence Interval
Z: Z-score (statistical test statistic))

## Author Contributions

A.O.A. and M.O.G. conceived and designed the study. M.O.G. and P.S.O. performed the laboratory procedures, while F.M.J., A.A., and A.O.A. conducted computational modeling and statistical analyses. M.O.G. drafted the initial manuscript, which was critically reviewed and revised by A.O.A. and A.A. A.O.A. supervised the project and managed administration. P.S.O. was responsible for sample selection and photomicrography. Data analysis and interpretation were carried out by A.O.A., F.M.J., and A.A. A.O.A. had full access to all study data and takes responsibility for the integrity and accuracy of the analyses. All authors contributed to the interpretation of results and approved the final version of the manuscript.

## Declarations

### Ethical Considerations

All relevant ethical guidelines were followed in the conduct of this study. The research did not involve direct patient participation; rather, it utilized de-identified histopathological samples prepared within institutional laboratory settings. Necessary approvals were obtained from the Health Review Ethical Committee of Lead City University, and all institutional protocols were observed. No clinical trial registration was required for this study.

## Acknowledgements

The authors gratefully acknowledge the invaluable support of Mr. Yinka Olaleye during the histopathological sample preparation and staining processes. We also extend our appreciation to the laboratory staff of the Department of Medical Laboratory Science, Lead City University, for their assistance in providing stains and other essential resources that contributed to the successful completion of this study.

